## Supplementary material for "Effectiveness and Challenges of Digital Tools Implementation for Enhancing Infectious Disease Surveillance Data Quality in Low- and Middle-Income Countries: A Systematic Review Protocol": Prisma P

| Title:   Identification | 1a | Effectiveness and Challenges of Digital Tools Implementation for Enhancing Infectious Disease Surveillance Data Quality in Low- and Middle-Income Countries: A Systematic Review Protocol |
| --- | --- | --- |
| Update | 1b | This is an initial protocol |
| Registration | 2 | Registered in Prospero with registration ID- 1023840 |
| Authors:   Contact | 3a | Oluwatosin Olu-Abiodun  Department of Nursing, Crescent University,  Abeokuta, Ogun State, Nigeria   Aderinsola Faturoti,  Department of Community Medicine,  Babcock University Teaching Hospital,  Ilishan-Remo, Ogun State, Nigeria ;  Akinmade Adepoju,  Department of Community Medicine,  Babcock University Teaching Hospital,  Ilishan-Remo, Ogun State, Nigeria ;  Davies Adeloye,  School of Health and Life Sciences, Nursing &Midwifery,  Teesside University, United Kingdom.   Akindele Adebiyi,  Department of Community Medicine and Public Health  University of Ibadan, Oyo state, Nigeria. ;  Olumide Abiodun,  Professor of Community medicine and Public Health  Babcock University Teaching Hospital, Ilishan-Remo, Ogun State, Nigeria.   Physical mailing address of the corresponding author: Department of Community Medicine, Babcock University Teaching Hospital, Ilishan Remo, Ogun State, Nigeria. |
| Contributions | 3b | Oluwatosin Olu-Abiodun- Conceptualisation, Protocol Draft and Review  Aderinsola Faturoti- Data collection and analysis  Akinmade Adepoju- Data collection and analysis  Davies Adeloye-Conceptualisation, Protocol Draft  Akindele Adebiyi-Protocol Draft and Review  Olumide Abiodun- Guarantor |
| Amendments | 4 | Any major changes to our original plan, such as adjustments in what studies to include, what outcomes we are interested in, or how we analyse the results, will be documented and shared on PROSPERO. |
| Support:   Sources  Sponsor | 5a  5b | None  None  None |
| Role of sponsor or funder | 5c | Roles of funder(s)- Not applicable  Roles of sponsor(s)- Not applicable  Roles of institution(s)- Not applicable |
| INTRODUCTION | | |
| Rationale | 6 | High-quality surveillance data are critical for mounting timely and effective outbreak responses, especially in Low- and Middle-Income Countries (LMICs). Digital surveillance tools enhance data collection, reporting, and analysis for optimising data quality. Despite the significant promise held by these digital platforms, their effectiveness in enhancing the quality of infectious disease surveillance data remains unclear, necessitating a critical and systematic inquiry.  The current evidence on the effectiveness of infectious disease digital surveillance tools for improving data quality in LMICs is mixed,  Considering the divergence in the evidence, a systematic evaluation is necessary to interrogate the effectiveness of digital surveillance platforms on infectious disease data quality in LMICs. This systematic review will consolidate available evidence to determine the overall impact of digital surveillance tools on infectious disease data quality while also identifying the contextual factors that influence the successful and unsuccessful implementation of digital surveillance. Furthermore, the review will provide policy and strategy recommendations to optimise digital surveillance for infectious diseases in LMICs. |
| Objectives | 7 | 1. To quantitatively and qualitatively assess the impacts of digital surveillance tools compared to conventional surveillance methods on data quality dimensions: timeliness, accuracy, completeness, and reliability. 2. To explore contextual and implementation factors (e.g., infrastructure, workforce capacity, data security, interoperability) influencing the effectiveness of digital surveillance tools. 3. To provide evidence-based recommendations for public health professionals, researchers, and policymakers on deploying digital tools in infectious disease surveillance. |
| METHODS | | |
| Eligibility criteria | 8 | **Study designs:** Eligible studies will include randomised controlled trials, quasi-experimental designs, cohort and case-control studies, cross-sectional analyses, program evaluations, and mixed-methods studies explicitly evaluating digital surveillance interventions.  **Case definitions:** Studies must clearly define infectious diseases monitored, adhering to internationally recognised or national surveillance case definitions. LMICs will be defined according to the World Bank's 2024 classification.  **Participants:** Our study participants are health systems, healthcare facilities, or public health agencies operating within low—and middle-income countries (LMICs) that use or assess surveillance systems for detecting and responding to infectious diseases.  **Interventions:** These include digital tools or platforms such as mobile applications, electronic reporting systems, web-based dashboards, and integrated health information technologies.  **Comparators:** Encompasses traditional surveillance methods (such as paper-based or non-digital approaches), no surveillance system, or pre-intervention periods in before-and-after studies (comparing baseline data with data collected following digital tool implementation).  **Outcomes:** Primary outcomes include timeliness, accuracy, completeness, and reliability of data. Secondary outcomes cover user acceptability, usability, adoption rates, implementation costs, and feasibility metrics.  **Timeframe:** studies published in English between January 2000 to April 2025 will be assessed over a duration of eight months. |
| Information sources | 9 | A systematic search will be conducted in five electronic databases: PubMed/MEDLINE, EMBASE, Global Health (CAB Abstracts), and CINAHL, complemented by searches in Google Scholar for relevant grey literature. Searches will be restricted to English-language studies published between January 2000 and April 2025. |
| Search strategy | 10 | The search strategy will combine keywords and Medical Subject Headings (MeSH) related to digital surveillance tools (e.g., "mobile health", "digital reporting systems", "electronic surveillance"), infectious diseases (e.g., "malaria", "cholera", "COVID-19"), and LMIC settings (country-specific terms alongside "low-income" or "middle-income"). |
| Study records:   Data management | 11a | The EndNote X8.2 (Bld 11343) software will be used to manage the records of the retrieved studies. |
| Selection process | 11b | An initial titles and abstracts screening will be undertaken to exclude irrelevant studies. All the remaining studies will have their titles and abstracts independently assessed for eligibility by two authors (AA and FA). The full articles of the abstracts and titles for which eligibility is not agreed on or remains unclear will be retrieved and jointly examined by both assessors (AA and FA) in conjunction with AO to engender a consensus, as required. Our study will maintain a record of all excluded studies and the reasons for exclusion. |
| Data collection process | 11c | Two reviewers will independently extract data using a standardised extraction form capturing bibliographic details, study design and duration, participant characteristics, intervention specifics, comparator methods, outcome definitions and measures, contextual implementation factors, and quantitative and qualitative outcomes.  Discrepancies will be resolved through consensus or arbitration by a third reviewer. Study authors will be contacted through email to provide unavailable data. |
| Data items | 12 | For each study, we will document bibliographic details (author, publication year, title, source) country of study, study design (cross-sectional, cohort, randomized controlled trial, mixed methods), study setting (hospital-based, community-based, national surveillance system), and target population (healthcare workers, surveillance officers, general).  The study will also document the study objectives and questions, sample size, and eligibility criteria. |
| Outcomes and prioritization | 13 | The study will extract data on the primary outcomes such as timeliness, accuracy, completeness, and reliability of data. Secondary outcomes cover user acceptability, usability, adoption rates, implementation costs, and feasibility metrics and their measurement methods. Secondary data to be extracted are the impact on disease detection, outbreak response efficiency, and intervention integration within the broader health information systems |
| Risk of bias in individual studies | 14 | We will utilise ROBINS-I for quality appraisal and tailor it to different study designs. For cohort studies, all seven domains remain relevant, with particular emphasis on confounding and selection bias due to the importance of temporal relationships.  In contrast, for cross-sectional studies, issues related to confounding, participant selection, and outcome measurement continue to be pertinent; however, domains concerning intervention classification and deviations from the planned interventions may be less applicable or may require reinterpretation based on how exposure is defined. Customising each domain involves explicitly defining what constitutes the 'intervention' as the key exposure and contextualising bias assessments within the specific temporal and structural aspects characteristic of each study design.  The assessment will be conducted by two independent assessors, while discrepancies will be resolved through a consensus-building process that may involve a third assessor.  Studies will be categorised by risk of bias as low, moderate, or high. |
| Data synthesis | 15a | The approach to the synthesis of findings from the included studies will be systematic and structured using quantitative and qualitative methods.  Quantitative synthesis will involve random-effects meta-analysis when comparable data are sufficient.  Qualitative thematic analysis will investigate implementation factors affecting tool effectiveness.  Mixed-methods integration will triangulate quantitative and qualitative findings, providing comprehensive insights.  All analyses will adhere to PRISMA 2020 guidelines and be conducted using Stata and Atlas-ti software. |
|  | 15b | Heterogeneity will be assessed using the I² statistic, with subgroup and meta-regression analyses exploring variability sources. |
|  | 15c | Our goal with this meta-analysis is to bring together and compare findings from various studies on how digital surveillance tools perform in low- and middle-income countries (LMICS). We will focus on the key outcome areas:   1. Primary outcomes: Data quality indicators like accuracy, completeness, timeliness, and reliability; how user-friendly the tools are; and the rates at which they are adopted. 2. Secondary outcomes: Digital platform usability and adoption rates, and how these tools influence outbreak detection and response, such as how early outbreaks are identified and whether reporting delays decrease. |
| Meta-bias(es) | 16 | We will construct a funnel plot to detect small-study effects or selective publication.  For meta-analyses of at least 10 studies, Egger’s regression test will be used to detect publication bias. If there are fewer than ten studies, we will use the method outlined in the Cochrane Handbook.  In the case that publication bias exists, a trim-and-fill method will be used to adjust for possible missing data and estimate the corrected effect size |
| Confidence in cumulative evidence | 17 | We will assess the certainty of evidence for each primary and secondary outcome using the GRADE approach, which considers five domains: risk of bias, inconsistency, indirectness, imprecision, and publication bias. The overall certainty will be rated as high, moderate, low, or very low. |
